# Social media addiction and its psychological effects among school students: A cross-sectional study from Kathmandu, Nepal

**DOI:** 10.1101/2025.07.08.25331070

**Authors:** Preeti Bhattarai, Dilip Roka Magar, Bharat Kafle, Pratik Bhattarai, Umesh Raj Aryal, Seshananda Sanjel

## Abstract

**Background:** Social media has become an integral part of daily activities. It has both positive and negative impacts on individuals. Using social media websites is one of the most common activities among today’s children and adolescents. Therefore, this cross-sectional, school-based study was conducted to assess the psychological effects of social media on adolescents in schools in Kathmandu city, Nepal.

**Methods:** A school-based cross-sectional study with a sample size of 215 was conducted to collect data. Ethical approval was obtained from the Institutional Review Committee of Karnali Academy of Health Sciences. Both informed assent and consent were obtained from all participants and their guardians. Out of seven government schools, four were selected using simple random sampling, and in the selected schools, a complete enumeration of grade 9 and 10 students was carried out. Two sets of standardized self-administered, semi-structured questionnaires—the Social Media Addiction Scale (SMASSF) and the Short Mood and Feelings Questionnaire (SMFQ)—were used. Data was entered into EpiData 3.1 and analyzed using IBM SPSS version 26.

**Results:** Approximately 51.20% of individuals with social media addiction were depressed, while 48.80% were not. Logistic regression analysis was performed to assess the association between the dependent variables. Followers of the Hindu religion were found to have 126% higher odds of being addicted to social media [OR = 2.259, 95% CI: (1.167–4.371)], and those living in nuclear families had 56% lower odds of social media addiction [OR = 0.438, 95% CI: (0.230–0.834)]. Students who owned their own phones had 138% higher odds of social media addiction [OR = 2.379, 95% CI: (1.177–4.808)]. Similarly, students who reported the availability of Wi-Fi at home had 231% higher odds of social media addiction [OR = 3.312, 95% CI: (1.526–7.187)].

**Conclusions:** This study highlights a significant association between social media use and depression. Factors such as family type, religion, hours of use, owning a mobile phone, and Wi-Fi availability at home are associated with depression. It is recommended to conduct longitudinal studies for further in-depth exploration of this life course, with a focus on early diagnosis, proper counseling, and providing love and support from friends and family to detect and address depression.

## Introduction

Social media refers to a wide range of applications, websites, and blogs that allow people around the world to connect via the internet, engage in chat, share content, make video calls, and enjoy many other features it offers to its users (1). Social media has recently become a part of people’s daily activities, as they spend a lot of time on platforms like Facebook, Messenger, Instagram, and other popular networks. This type of media is particularly popular among the younger generation (2–3). Social media has both positive and negative impacts on individuals. Its overuse can lead to serious health problems like depression, anxiety, mania, eating disorders, sleep deprivation, insecurity, FOMO (Fear of Missing Out), internet addiction, and other antisocial behaviors. The use of social media has developed exponentially ever since the rise of the internet (4). Social media addiction is a subcategory of internet addiction (5). Recent global survey 2020 by Statista, 34% admitted to social media addiction among the internet users. Similarly, a systematic review and Meta analysis showed the complex relationship between social media and adolescent psychological development. Excessive use of social media leads to cyber bullying and this negative effect can improve by social support from both peers and parents (6).

A scoping review showed a significant 95% increase in smart phone use among adolescents, with 46% using their devices for more than three hours daily. The results indicate that using social media and owning a smart phone are the most common activities among today’s children and adolescents. The increasing psychological effects due to social media addiction remain an iceberg phenomenon, with much of the impact still hidden or unaddressed (7).

Kathmandu revealed that the prevalence of social media addiction ranges from 29.9% to 46 % (8–11). The results varied slightly in different parts of Nepal, with a prevalence of 13.3% to 51.2% (12–14). Among those addicted to social media, the reported psychological effects range from 21% to 68.2% (8–10). Adolescents, in particular, are susceptible to the negative effects of social media due to their transitional phase from childhood to adulthood. Their emotional vulnerability and sensitivity often magnify minor issues, impacting their psychological well-being. Key areas of concern related to adolescent social media use include cyberbullying, exposure to inappropriate content, sleep deprivation, internet addiction, "Facebook depression," and social isolation (6, 13). According to the Nepal Social Media Users Survey 2021, two in five users spend 1-2 hours on social media every day, while 39% spend between 3-6 hours daily. Additionally, 92% of users engage with social media during their leisure time. This implies that the use of social media is common among Nepalese adolescents, which poses a threat of various psychological effects on this group due to their vulnerability (15). Nepal has adopted various cyber security laws, which are often referred to as laws for the internet (16). Although these laws do not encompass the psychological effects of social media addiction, some initiatives have been taken by the non-governmental sector to address these effects, which are often not satisfactory. The national health system and school education have not yet incorporated mental health interventions to address this emerging public health problem (17). It is high time to tackle this burden and explore the issues that have not been sufficiently covered. In line with this, we need to fill the knowledge gaps at the school level and raise awareness from the local level to the federal government. Therefore, this study was conducted to assess the psychological effects of social media among adolescents in grades 9 and 10 in schools in Kathmandu city, Nepal.

## Materials and Methods

Present study followed the STROBE (Strengthening the Reporting of Observational Studies in Epidemiology) guidelines for reporting purposes (18).

### Study design and settings

A cross-sectional study was conducted to collect data from a representative sample of students in a school setting. The research was carried out in government schools within Tokha Municipality of Kathmandu District. According to the education section staff of Tokha Municipality, there are nine government schools, seven of which are secondary schools, and 81 private schools listed in the municipality.

### Sample size and sampling

The sample size was calculated based on 46% prevalence of social media addiction i.e., p=0.46 and q=0.54 (8) using the following formula: 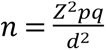 (19), where d (allowable error) is 7%. With the addition of a 10% non-response rate, the calculated sample size was 214.74, rounded to 215. Out of the seven government schools in Tokha Municipality, four were selected using simple random sampling. The sampling flow diagram of the study presented below (Fig 1).

**Figure 1:**
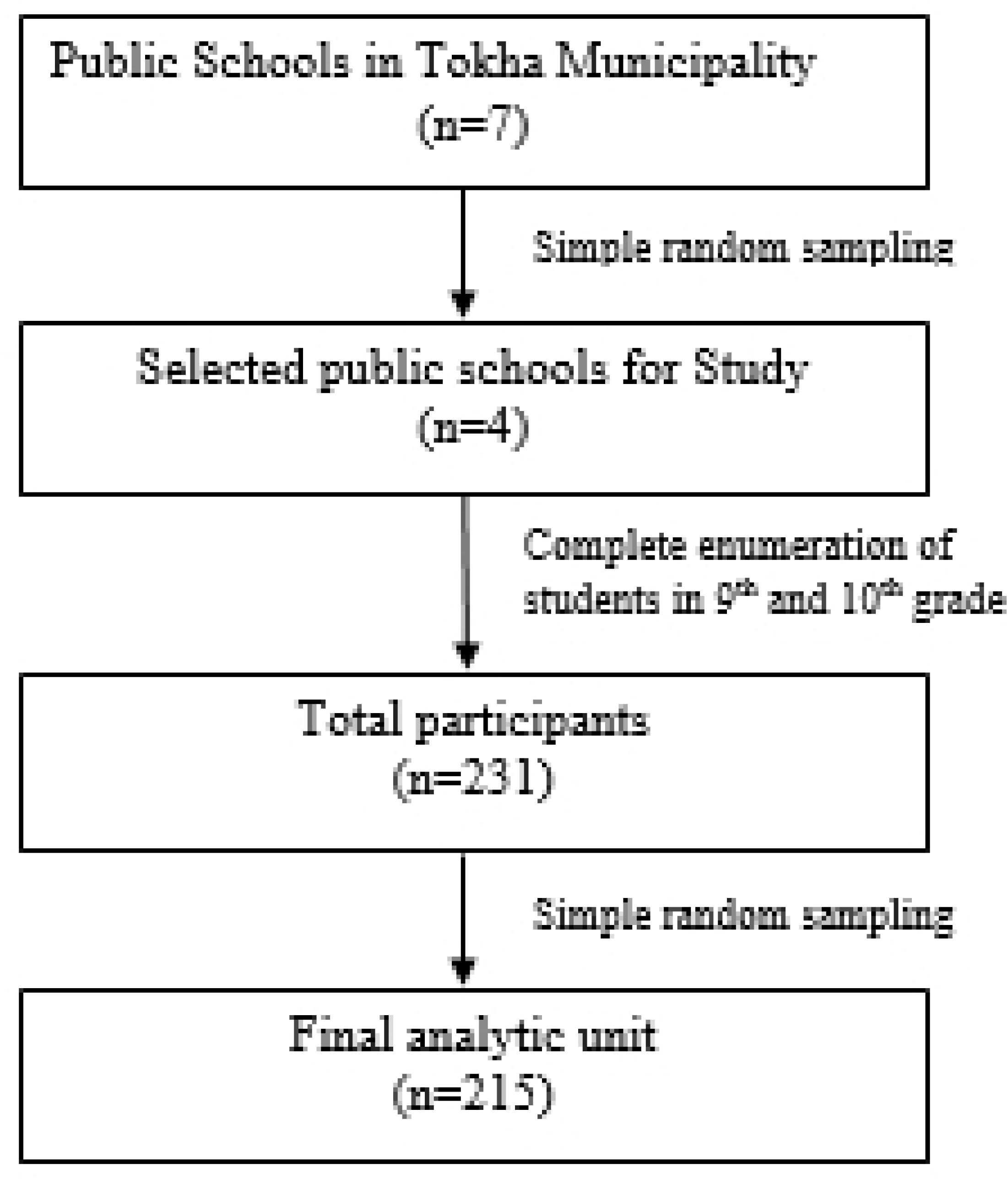
Steps for selecting schools and students.

### Data collection

Study was conducted between 5^th^ September, 2023 to 10^th^ September, 2023, data were collected from Grade 9 and 10 students at four selected public schools. Two sets of standardized, self-administered, semi-structured questionnaires—the Social Media Addiction Scale for Students (SMASSF) (20) and the Short Mood and Feelings Questionnaire (SMFQ) (21)—were adopted.

### Study validity and reliability

Validity of the study were assessed by used self-administered questionnaire of 29 questions with 5-point Likert-type scale adopted from earlier study of Nepal (8, 20, 21). Translation and back translation of tools (English-Nepali) was performed, peer-reviewed, expert consultation, and pre-testing were conducted. The questionnaires were reviewed to fit the Nepali context (8). Pretesting was conducted, and the tools were finalized before data collection to maintained validity whereas reliability of questionnaire was determined by internal consistency tested across 29 questions with 5-point Likert-type scale twenty question items used in this study with Cronbach coefficient alpha score of 0.91, which indicated high reliability of this tool (22). Parallel test statistically significant with <0.001 p-value at 433 degree of freedom that statistically signifies the tool used was fit for the study purposed. Similarly, reliability of Short Mood and Feelings Questionnaire was adopted (8,20,21) and determined by internal consistency tested across 13 questions, with responses ranked as: Not true, Sometimes, and True questions 3-point Likert-type scale items used in this study with Cronbach coefficient alpha score of 0.88, which indicated high reliability of this tool (22). Parallel test statistically significant with <0.001 p-value at 89 degree of freedom that statistically signifies the tool used was fit for the study purposed.

### Study variables

#### Outcome

The dependent variable was measured using the SMASSF tool, which includes a 5-point Likert-type scale consisting of 29 items across 4 sub-dimensions: items 1–5 fall under the virtual tolerance sub-dimension; items 6–14 fall under the virtual communication sub-dimension; items 15–23 fall under the virtual problem sub-dimension; and items 24–29 fall under the virtual information sub-dimension. The scale includes 3 positive and 2 negative items, and it uses a 5-point grading system. The highest possible score on the scale is 145, while the lowest is 29. Data showed a positive skew during the normality test, with a skewness score of 0.842 (S.E. 0.166). Thus, in this study, the dependent variable was dichotomized using the median as a cutoff value, with scores below the median considered "not social media addicts" and scores above the median considered "social media addicts." The median value was 76, with higher scores indicating social media addiction.

### Predictors

#### Socio-demographic factors

Age, Gender, grade, religion, and type of family were used. Age was categorized into dichotomous variable which defined by “age between 10-16 years categorized early adolescent and 17-19 years age categorized late adolescent”. Gender was categorized either “male” or “female” and grade also defined as either “grade 9” or “grade 10”. Religion was categorized into either “Hindu” or “Non-Hindu”. Type of family categorized into “nuclear” and “joint”.

#### Socio-economic factors

Family income, father occupation, and mother occupation were used as a socio-economic factor. Family income was categorized into dichotomous analysis of median value which was 30000, further categorized into “above 30000” and “below 30000”. Father occupation was categorized into “business, agriculture, and service”. Mother occupation was categorized into “house-maker, business, service, and agriculture”.

#### Access and availability of social media related factors

An hour use of social media, Wi-Fi availability at home, mobile phone ownership, and preference time of social media use were used. An hour use of social media was categorized into “less than one hour and more than one hour”. Wi-Fi availability at home categorized either “yes” or “no” and owning mobile phone also categorized into “yes” and “no”. Preferable time for social media used was categorized into “morning, evening, and night”.

#### Psychological effect factor

The Short Mood and Feelings Questionnaire (SMFQ) were used as a screening tool to assess suspected depression among school-going adolescents. It consists of 13 questions, with responses ranked as: Not true (score = 0), Sometimes (score = 1), and True (score = 2). Data showed a positive skew during the normality test, with a skewness score of 0.544. In this study, the independent variable was dichotomized using the median as a cutoff value, with scores below the median considered "not suspected depression" and scores above the median considered "suspected depression." The median value was 7, with higher scores indicating suspected depression.

### Data analysis

Data were entered into EpiData 3.1 and analyzed using IBM SPSS version 26. Normality, multicollinearity, mean, standard deviation, and descriptive analyses were performed. For inferential analysis, chi-square tests, binary logistic regression, and multivariate logistic regression were conducted to assess goodness of fit and the association with the dependent variable. In the final adjusted model, multicollinearity was observed, with all Variance Inflation Factors (VIF) below two. Table 1 and 2 shows the mean, standard deviation, and descriptive analyses of categorical variables which presented the frequencies and percentages. Fig 2 shows the prevalence of the study. Similarly, table 3 shows the bivariate relationships between dependent and independent variables were assessed using the Pearson Chi-square test of independence. The best-fitting model, determined by excluded one multicollinearity variable income. Therefore, the final adjusted model (Table 4) included Depression, Income level, Religion, Screening time, Preference time, Grade, Owning phone status, and Wi-Fi availability at home. Furthermore, both unadjusted and adjusted multivariable logistic regression models were conducted to explore the factors associated with social media addiction. Odds ratios (OR) and 95% confidence intervals (CI) from both unadjusted and adjusted multivariable logistic regression models values of OR below 1 represent low level or protective level of media addiction and above 1 OR indicate the odds of experiencing media addiction at different levels of the independent variables are presented same table. And statistical significance was defined as a p-value less than 0.05.

**Figure 2:**
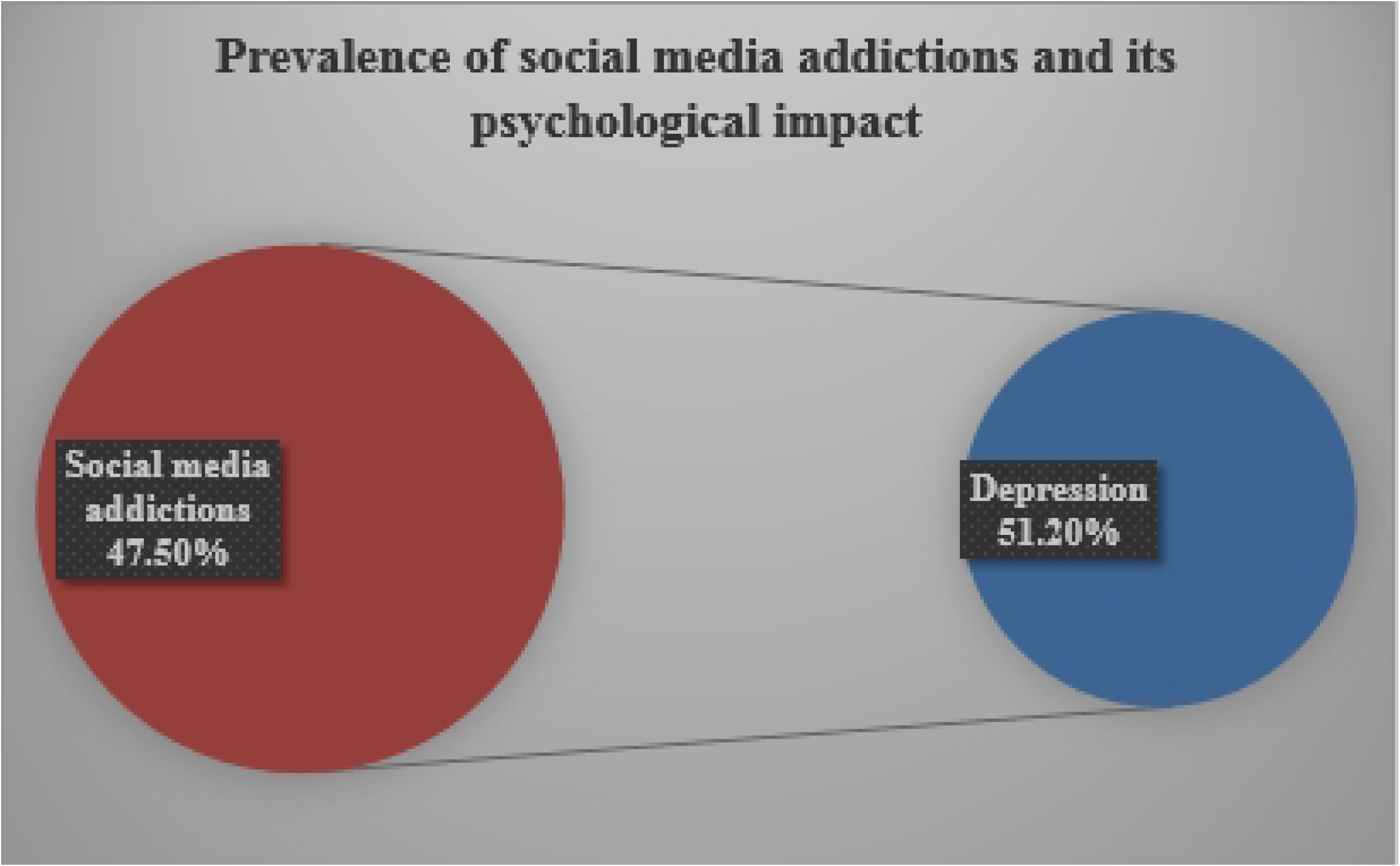
Social Media Addiction and Its Psychological Impact.

**Table 1:** Socio-demographic characteristics of the participants n=215.

| Variables(n=215) | Frequency(n) | Percentage (%) |
| --- | --- | --- |
| **Age category (15.30±1.14)** |  |  |
| Early adolescent | 184 | 85.6 |
| Late adolescent | 31 | 14.4 |
| **Grade** |  |  |
| 9 | 129 | 55.3 |
| 10 | 96 | 44.7 |
| **Gender** |  |  |
| Male | 103 | 47.9 |
| Female | 112 | 52.1 |
| **Religion** |  |  |
| Hindu | 142 | 66 |
| Non-Hindu | 73 | 34 |
| **Family** |  |  |
| Nuclear | 128 | 59.5 |
| Joint | 87 | 40.5 |
| **Category of income** |  |  |
| Below 30000 | 125 | 58.1 |
| Above 30000 | 90 | 41.9 |
| **Father's occupation** |  |  |
| Business | 37 | 17.2 |
| Agriculture | 91 | 42.3 |
| Service | 87 | 40.5 |
| **Mother's occupation** |  |  |
| House-maker | 137 | 63.7 |
| Business | 37 | 17.2 |
| Service | 41 | 19.1 |
| Agriculture | 91 | 42.3 |

### Ethical consideration

The Institutional Review Committee at Karnali Academy of Health Sciences, Jumla, Nepal provided the ethical permission (reference number 080/081/4) in August 2023. Written Informed consent and assent were obtained from all participants and their guardians. Students from grades 9 and 10 were selected from government schools. Administrative approvals were obtained from all selected school with reference numbers 11/2080/081, 1359/2090/081, 19/2080/081, and 22/2080/081.

## Results

### Socio-demographic characteristics

Table 1 below shows the demographic characteristics of a total of 215 participants took part in the study. The mean age of the participants was 15.30 ± 1.14 years. Approximately 85.6% of the participants were early adolescents, and 55.3% were in grade 9. More than half (52.1%) of the participants were female, and 66% were Hindu. The majority (59.5%) of the participants came from nuclear families, and 58.1% had an average monthly income below 30,000. Most of the participants’ fathers were engaged in agriculture (42.3%), followed by service (40.5%) and business (17.2%).

### Variables related to availability of internet facility

Table 2 below shows the availability of internet facility among the 215 participants, 70.7% owned a phone, of which 77.7% had Wi-Fi access. Additionally, 66.5% used Wi-Fi for less than an hour, with their preferred time of usage being at night (54.4%), followed by the evening (26.5%) and morning (19.1%).

**Table 2:** Variables related to availability of internet facility n=215.

| **Variables(n=215)** | **Frequency(n)** | **Percentage (%)** |
| --- | --- | --- |
| **Owning phone** |  |  |
| Yes | 152 | 70.7 |
| No | 63 | 29.3 |
| **Wifi availability at home** |  |  |
| Yes | 167 | 77.7 |
| No | 48 | 22.3 |
| **Hour of used** |  |  |
| Less than 1 hour | 143 | 66.5 |
| More than 1 hour | 72 | 33.5 |
| **Preferable time used** |  |  |
| Morning | 41 | 19.1 |
| Evening | 57 | 26.5 |
| Night | 117 | 54.4 |

### Association between variables and social media addiction scale

Table 3 below shows that the factors associated with social media addiction are significantly correlated with depression (p = 0.002), religion (p = 0.004), type of family (p = 0.018), hours of use (p = 0.018), ownership of a mobile phone (p = 0.001), and Wi-Fi availability at home (p = 0.001).

**Table 3:**
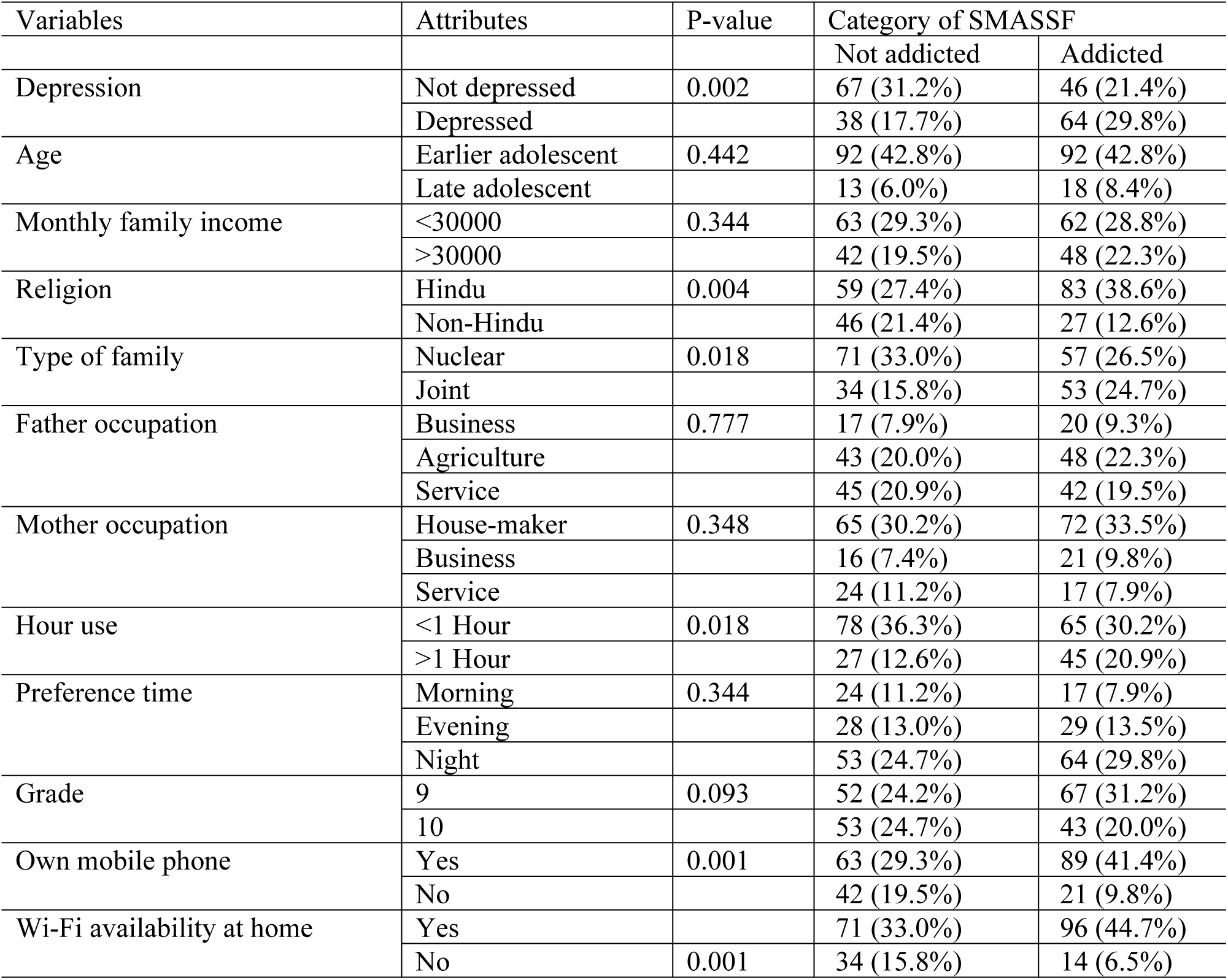
Association between variables and social media addiction scale n=215.

### Prevalence of social media addiction and its psychological impact

About 51.20% of social media addictions were depressed and 48.80% of the people who were social media addiction were not depressed (Fig 2).

### Multiple logistic regression analysis of social media addiction scale (SMASSF)

Table 4 below presents the logistic model results. In the unadjusted model, the factors of depression, Hindu religion, nuclear family structure, usage of social media for less than one hour, ownership of a phone, and availability of Wi-Fi at home were found to be significant. The application of the Short Mood and Feelings Questionnaire indicated that individuals who were not depressed had 63% lower odds of social media addiction [OR = 0.370, 95% CI: (0.199-0.689)]. Among various sociodemographic variables, religion and type of family were significant in the final adjusted model. Followers of the Hindu religion had 126% higher odds of being social media addicts [OR = 2.259, 95% CI: (1.167-4.371)], while those living in nuclear families had 56% lower odds of social media addiction [OR = 0.438, 95% CI: (0.230-0.834)]. Students who owned their own phones had 138% higher odds of social media addiction [OR = 2.379, 95% CI: (1.177-4.808)]. Similarly, students who reported having Wi-Fi availability at home had 231% higher odds of social media addiction [OR = 3.312, 95% CI: (1.526-7.187)].

**Table 4:** Multiple logistic regression analysis and social media addiction (SMASSF) n=215.

| Variables | 95% C.I. for (Lower- Upper Bound) |  |
| --- | --- | --- |
|  | ORu | ORa |
| **Ref (Depressed)** |  |  |
| Not depressed | 0.408 (0.235-0.706) | 0.370 (0.199-0.689) |
| **Ref (>30000)** |  |  |
| <30000 | 0.861(0.501-1.482) |  |
| **Ref (Non-Hindu)** |  |  |
| Hindu | 2.397(1.341-4.284) | 2.259 (1.167-4.371) |
| **Ref (Joint family)** |  |  |
| Nuclear family | 0.515(0.296-0.896) | 0.438(0.230-0.834) |
| **Ref (>1 Hour)** |  |  |
| < 1 Hour | 0.500(0.280-0.893) | 0.572(0.288-1.134) |
| **Ref (Preference time in night)** |  |  |
| Morning | 0.587(0.285-1.205) | 0.786(0.342-1.807) |
| Evening | 0.858(0.455-1.617) | 0.895(0.425-1.885) |
| **Ref (Grade 10)** |  |  |
| Grade 9 | 1.588(0.924-2.729) | 1.415(0.756-2.648) |
| **Ref (No owning of phone)** |  |  |
| Owning Phone | 2.825(1.527-5.227) | 2.379(1.177-4.808) |
| **Ref (Wi-Fi not available at home)** |  |  |
| Wi-Fi available at home | 3.284(1.641-6.572) | 3.312(1.526-7.187) |

## Discussion

The study examined social media addiction and its psychological effects among school-going adolescents in grades 9 and 10. The results showed that 51.20% of the adolescents were addicted to social media, while 58.2% of those who were addicted reported experiencing depression. Additionally, the study found that social media addiction is significantly associated with depression, religion, type of family, hours spent using social media, ownership of a mobile phone, and Wi-Fi availability at home.

The study conducted in Nepal showed that social media addiction ranges from 35.4% to 46% (8–10). Various studies reporting on social media addiction and its psychological effects in Nepal indicate prevalence rates ranging from 21% to 68.2% (8–10). Studies conducted in Europe, America, and Asia show prevalence rates ranging from 18.4% to 36% (23–25). In contrast, studies conducted in Asia and Southeast Asia report similar levels of prevalence, ranging from 34.4% to 46.4%. This similarity may be attributed to comparable socioeconomic conditions, cultural factors, and belief systems in this region (26, 27). In line with these findings, the present study indicates that social media addiction is significantly lower among those who are not depressed. This result is consistent with various studies conducted in India, Austria, and Ethiopia (28–30). Excessive time spent on social media can lead to addiction and reduced face-to-face interactions and real-world connections, which are crucial for mental health. This isolation from social cohesion and focus solely on virtual worlds ultimately pushes individuals toward depression.

Nepal has a large, youthful Hindu population, and the younger generation is more prone to social media use. Many young Hindus engage with social media, and in recent years, an increasing number of people have used these platforms to express and promote their religious and cultural identities. Hindu festivals, practices, and pride in one’s religious heritage are often shared online. This increased engagement could lead to more time spent on these platforms to connect with similar communities or participate in religious discourse. As a result, followers of the Hindu religion have 126% higher odds of social media addiction, a finding that is consistent with studies conducted in Kathmandu (31).

In joint families, particularly where both parents are working or busy, children often interact within the household while their elder relatives are frequently engaged in social media. Consequently, children from these families spend a lot of time on social media, taking photos, uploading them, and discussing likes and shares. Similarly, those who need guidance for book reading and physical social interactions tend to engage in social media, which is imitated by both children and adolescents. They often become familiar with upgraded versions of social media platforms, requiring even more time, which can lead to addiction. The study demonstrated that nuclear families have 56% lower odds of social media addiction, a result that aligns with various studies conducted in Pakistan but contrasts with studies conducted in India and Nepal (32–34).

Individuals who spend excessive amounts of time on social media may begin to show signs of addiction, which is commonly associated with factors related to social media addiction in Bangladesh, the UK, Sweden, India, and Kathmandu (10, 35–38). Despite this, the results of the present study contrast with previous findings from the final adjusted model.

The present study revealed that individuals who owned their own phones had 138% higher odds of social media addiction. This finding is consistent with previous studies conducted in Kathmandu, Africa, and Saudi Arabia (8, 39, 40). This may be due to the fact that easy access to mobile phones makes it more tempting to check apps frequently throughout the day, leading to habitual use and, over time, addiction.

Students who reported the availability of Wi-Fi at home had 231% higher odds of social media addiction. This result is consistent with various studies conducted in Nigeria, Nepal, Pokhara, and Myagdi, Nepal (34, 41–43). This may be due to the fact that the availability of Wi-Fi allows individuals to spend unlimited time on social media, thanks to constant access and convenience for various purposes such as studying, entertainment, gaming, and social media use.

### Limitation and strengths

This study was conducted with a sample of adolescents based on self-reported questionnaires; therefore, the generalizability of these results may vary.

## Conclusions

Social media addiction is a major issue among adolescents in grades 9 and 10, with almost half of the respondents suffering from it. This study highlights the significant association between social media use and depression, as well as the correlation with factors such as type of family, religion, hours of use, ownership of a mobile phone, and Wi-Fi availability at home. It is recommended that longitudinal studies be conducted for further in-depth research on this life course to detect depression. Since it is treatable, early diagnosis, proper counseling, and support from friends and family can help. Additionally, there is a need to develop policies and programs to mitigate the negative psychological effects of excessive cell phone use, which can lead to social media addiction.

## Data Availability

All data produced in the present work are contained in the manuscript

## Acknowledgment

We would like to acknowledge Tokha municipality, all selected schools, student participants, parents and others who directly and indirectly contributed to the success of the study.

## Conflict of interest

None

## Funding

None

## Author Contributions

Conceptualization: Preeti Bhattarai, Dilip Roka Magar, Seshananda Sanjel

Data curation: Preeti Bhattarai, Dilip Roka Magar

Formal analysis: Preeti Bhattarai, Dilip Roka Magar, Umesh Raj Aryal, Bharat Kafle

Investigation: Dilip Roka Magar

Methodology: Preeti Bhattarai, Dilip Roka Magar, Bharat Kafle

Resources: Preeti Bhattarai, Dilip Roka Magar

Supervision: Preeti Bhattarai, Seshananda Sanjel Visualization: Bharat Kafle

Writing-original draft: Preeti Bhattarai, Pratik Bhattarai, Bharat Kafle,

Writing-review & editing: Preeti Bhattarai, Bharat Kafle, Pratik Bhattarai, Umesh Raj Aryal, Seshananda Sanjel

## Notes

### Competing Interest Statement

The authors have declared no competing interest.

### Funding Statement

This study did not receive any funding

### Author Declarations

Ethical approval was obtained from the Institutional Review Committee of Karnali Academy of Health Sciences (reference number 080/081/4) in August 2023.

